# Successive epidemic waves of cholera in South Sudan, 2014 - 2017

**DOI:** 10.1101/2020.10.09.20209262

**Authors:** Forrest K Jones, Joseph F Wamala, John Rumunu, Pinyi Nyimol Mawien, Kol Mathew Tut, Shirlee Wohl, Lul Deng, Lorenzo Pezzoli, Linda Haj Omar, Justin Lessler, Marie-Laure Quilici, Francisco J Luquero, Andrew S Azman

**Affiliations:** Department of Epidemiology, Johns Hopkins Bloomberg School of Public Health, Baltimore, USA; World Health Organization, Juba, South Sudan; South Sudan Ministry of Health, Juba, South Sudan; World Health Organization, Geneva, Switzerland; World Health Organization, Brazzaville, Republic of Congo; Institut Pasteur, Paris, France; Epicentre, Geneva, Switzerland; Médecins Sans Frontières, Geneva, Switzerland

## Abstract

**Background:** Between 2014 and 2017, successive cholera epidemics occurred in South Sudan within the context of civil war, population displacement, flooding, and drought. Understanding the determinants of cholera spread in complex settings like this can provide valuable insights for mitigating future cholera risk.

**Methods:** We analyzed cholera linelist and molecular data to describe the spatio-temporal progression of the epidemics. We explored the role of rainfall, population movement and vaccination campaigns in shaping the explaining incidence and the spatial distribution of reported cases.

**Findings:** South Sudan experienced three distinct cholera epidemic waves of cholera ranging from 6-18 months with more than 28,000 cases reported and more than 2 million cholera vaccine doses delivered to curb transmission. The 2014 and 2015 epidemics remained spatially limited while the 2016/17 epidemic exploded along the Nile river. Initial cases of each epidemic were reported in or around Juba soon after the start of the rainy season, but we found no evidence that rainfall modulated transmission during each epidemic. All isolates analyzed had similar genotypic and phenotypic characteristics, closely related to sequences from Uganda and Democratic Republic of Congo. The direction of large-scale population movements between counties with cholera outbreaks was consistent with the spatial distribution of outbreaks. As of September 2020, zero cholera cases have been confirmed within South Sudan since 2017.

**Interpretation:** The three epidemic waves were caused by *V. cholerae* of the same clonal origin despite the periods of no reported cases between waves. While the complex emergency likely shaped some of the observed spatial and temporal patterns of reported cases, the full scope of transmission determinants remains unclear. Timely and well targeted use of cholera vaccine can avert cases and deaths, however, most of the vaccine campaigns occurred after the epidemic peak highlighting the challenges of delivering vaccines quickly in response to an outbreak in settings like South Sudan. These analyses provide a multi-faceted template for examining cholera dynamics through epidemiological, microbiological, climatic, and behavioral lenses.

**Funding:** *The* Bill and Melinda Gates Foundation

## Research in context

### Evidence before this study

We searched PubMed for articles published between January 1st, 1970 and September 18th, 2020 using “cholera”[title] AND (epidemic OR outbreak) AND (Africa OR emergency OR conflict) AND (molecular OR sequencing OR rain OR precipitation OR mobility OR movement OR vaccine OR WASH). We also downloaded reported cases and deaths from the Global Health Observatory Data repository on all outbreaks in the geographical area of South Sudan. Across the globe, there are an estimated 2.9 million cases and 95,000 deaths annually. Many of these cases occurred in areas with poor infrastructure including peri-urban slums, rural areas dependent on surface water, and areas with complex emergencies.

Cholera has been introduced into the African continent several times over the past 50 years. In particular, endemic transmission has been maintained in several countries bordering the African Great Lakes, including the Democratic Republic of Congo, Uganda, Tanzania, and Malawi. In contrast, reported cholera in South Sudan appears in cycles, with few reported outbreaks prior to 2005, a period of annual epidemics reported between 2005 and 2009, and then no reported cases between 2010 and 2014. The South Sudanese Civil War began in December 2013, leading to damaged essential infrastructure and displaced hundreds of thousands of individuals and a return of cholera, in successive annual waves starting in 2014. Previous studies have demonstrated associations between epidemic cholera and complex emergencies (e.g., Yemen, Haiti, Democratic Republic of the Congo), mass population movement (e.g., Senegal), and precipitation (e.g., Yemen and Haiti); however it is unclear how these factors interact to shape cholera epidemics. Oral cholera vaccination campaigns have only recently been used both in response to epidemics and preventatively thought its impact at the population level has not been well described.

### Added value of this study

Our study helps explain the complex story of three epidemic waves of cholera in South Sudan that began soon after the beginning of the civil war. We showed how each wave began in the capital city of Juba at the start of the rainy season and spread to different areas of the country. Using molecular data, isolates from all four years were found to be from the same clonal origin. We presented evidence, including the analyses of previously published whole genome sequences, that cholera was likely introduced in 2013 or 2014 from Uganda or the Democratic Republic of Congo and may have been re-introduced through multiple cross-border transmission events. We identified large-scale population displacement and movement as partially explaining the differences in the number of cases between years and transmission during the dry season of the 2016-2017 wave. Lastly, we describe 36 vaccination campaigns providing over 2 million doses of cholera vaccine: we find that early vaccination may have reduced local outbreak sizes though most vaccination activities occurred after the peak week of cases.

### Implications of all the available evidence

Our research highlights the need for regional-level responses to curb outbreaks of cholera in humanitarian settings. In addition to oral cholera vaccine campaigns and interventions to improve water, sanitation, and hygiene in these vulnerable settings, controlling cholera in nearby countries that have the potential to introduce cholera might be an effective additional strategy to reduce cases. Increased sampling of *Vibrio cholerae* from both human cases and environmental samples combined with whole genome sequencing can help us better understand how cholera spreads across borders and maintains itself in the environment. Expanding the use of sequencing and bioinformatics in more countries with *Vibrio cholerae* transmission could lead to improved global representativeness of sequences and integration of results into local decision-making processes. While increased precipitation has been linked to increased cholera transmission in many settings, we found that it was associated with the onset of outbreaks, but not with increases in transmission once an outbreak has begun. While our study adds to the evidence on the role of population movement in cholera outbreaks, improved methods for measuring population movement within and between countries during complex emergencies is needed to better characterize these relationships.

## Introduction

The seventh cholera pandemic, first recognized in 1960, continues to plague populations with poor access to safe water and sanitation across the world. Although estimates of the global burden are uncertain, over 140,000 suspected cholera cases are reported annually from Sub-Saharan Africa.^1^ Universal access to water and sanitation would likely eliminate cholera transmission but is unlikely given the current pace of progress and financial commitments.^2^

In 2017, the World Health Organization (WHO) led Global Task Force on Cholera Control (GTFCC) developed a roadmap to end cholera by 2030 and in 2018 a resolution to end cholera was adopted at the 71^st^ World Health Assembly.^3^ The roadmap calls for a geographically-targeted approach to use limited resources in areas with a high-risk of cholera, requiring an in-depth understanding of transmission across endemic and epidemic settings. South Sudan is one of the 47 focal countries of this roadmap. Detailed analyses of historical cholera in South Sudan, which has suffered from civil-war, mass population movements, floods and droughts, can provide important lessons for cholera control in similar epidemic-prone settings.

Three successive epidemic waves of cholera occurred in South Sudan between 2014 and 2017, leading to tens of thousands of cases and hundreds of deaths, many among internally displaced persons (IDPs).^4–6^ Each wave varied greatly by geographic extent, timing, and magnitude and were separated by periods, with few to no reported cases. It is unclear whether each outbreak was the result of a new introduction of *Vibrio cholerae* O1 or whether cholera was present but undetected between outbreaks. Here, using detailed cholera case data collected from 2014 through 2017, we describe the key features of the successive epidemic waves and explore how precipitation, population displacement, and vaccination campaigns may explain the differences between waves.

## Methods

### Cholera surveillance system

The Ministry of Health Republic of South Sudan (MoH) in collaboration with local health authorities implemented cholera surveillance through both the Integrated Disease Surveillance and Response (IDSR) and Early Warning Alert and Response Network (EWARN) with support from WHO and the Health Cluster. Linelist data were collected from all sites treating cholera patients across the country, including cholera treatment centers and oral rehydration points from June 2014 to December 2017. Prior to the confirmation cholera, a suspected cholera case was defined as any patient aged ≥5 years who developed severe dehydration or died from acute watery diarrhoea. After cholera was confirmed within a county, a suspected cholera case was any patient ≥2 years who developed acute watery diarrhoea. A confirmed case was a suspected case with culture-confirmed *V. cholerae* O1. We use the term epidemic waves to describe a rise and decline of reported cholera cases at the national-level, which is a different concept than the previously described ‘waves’ of the 7th pandemic ^7^.

### Laboratory analysis

When suspected cases were first reported in a county, stool specimens from select suspected cases were sent to the National Public Health Laboratory (NPHL) for culture confirmation.^8^ Additional isolates were collected from suspected cases and tested by culture at the NPHL in 2015 in a vaccine effectiveness study.^9^ Overall, 150 isolates from South Sudan (and 10 from Uganda) were sent to the French National Reference Center for Vibrios and Cholera (Institut Pasteur, Paris, France) for serotype, antibiotic resistance pattern testing (disk diffusion following Comité de l’Antibiogramme de la Société Française de Microbiologie 2013 standards for Enterobacteriaceae) and Etests (AB bioMérieux, Solna, Sweden) for the determination of minimum inhibitory concentrations (MICs) of nalidixic acid and ciprofloxacin), genotyping tests (*rstR, ctxB, tcpA, gyrA, gyrB, parE, parC*),^10^ and multi-locus variable number tandem repeat analysis (MLVA).^8,11,12^

### Rainfall data

Given that few sites directly measured rainfall in South Sudan during the study period, we used remote sensing-derived estimates. We obtained average daily precipitation estimates from 2010-2018 at the country, state, and county level from the Climate Hazards Group InfraRed Precipitation with Station Data dataset (https://climateserv.servirglobal.net/).

### Population displacement data

We used county-level population estimates from WorldPop (https://data.humdata.org/organization/worldpop). To better understand population movements as a result of the conflict, we acquired Mobility Tracking Data from the International Office of Migration (IOM) (https://www.globaldtm.info/). During the 2nd Round of Mobility Tracking (Mar-Apr 2018), IOM officers interviewed key informants (e.g. local authorities, community leaders and humanitarian partners) in 46/78 counties to estimate the number of individuals (internally displaced persons and returnees) present at assessment areas. These counties include 18/29 cholera-affected counties accounting for 62% of suspected cholera cases across the study period. Key informants also provided information on the year when individuals arrived and from where they came.^13^

### Intervention data

WHO collated data from all oral cholera vaccination (OCV) campaigns conducted between 2014 and 2018. We attempted to collate data on water, sanitation and hygiene (WASH) interventions but these data were not systematically reported.

### Data Analysis

We calculated the weekly number of cases, attack rate (AR), case-fatality ratio (CFR), the proportion of cases <5 years old, and proportion male. For most cases, we relied on the self-reported date of symptom onset for each case; when this was not available, we used the date of health facility visit. Administrative vaccination coverage was calculated by dividing the total number of first round doses delivered by the estimated target population size. To explore the potential effect of vaccination campaigns, we compared the attack rates of outbreaks where vaccine was used before the epidemic peak with those where it was used after the peak with Poisson regression models using the population size as an offset.

We generated a maximum likelihood tree from the 1210 publicly available *V. cholerae* O1 whole genomes.^14–16^ This tree closely resembles the phylogeny of 1203 genomes from Weill et al,^14^ but includes 7 genomes from Uganda published separately.^16^ GenBank accession numbers for all 1210 sequences are available in the supplement.^17^ For each reference-based assembly (reference accession:AE003852/AE003853), we masked recombinant sites manually (https://figshare.com/s/d6c1c6f02eac0c9c871e) and using Gubbins v2.3.4^18^ as previously described.^15^ We generated the tree from the ‘filtered_polymorphic_sites’ FASTA outputted by Gubbins, which contained only the 10,098 variant sites in our alignment. We used IQ-TREE v1.6.10 ^19^ with a GTR substitution model and 1000 bootstrap iterations.^20^ We rooted the tree on A6 by first constructing a tree containing an outgroup sequence (M66,accession:CP001233/CP001234) and choosing the most ancestral non-outgroup strain. We visualized phylogenetic trees using FigTree v1.4.4 (http://tree.bio.ed.ac.uk/software/figtree/). For MLVA analysis, we defined a clonal complex as a group of isolates where each differs (in loci-specific copy number) from at least one other isolate in the complex at no more than one locus.

We used a distributed lag non-linear model^21–23^ to investigate how accumulated rainfall over the previous seven days (AR7D) is associated with changes in cholera transmission in Juba County, as characterized by the instantaneous basic reproductive number, R_0_(t). We first jointly estimated the serial interval and reproductive number for the period of epidemic growth for each wave using methods described by White and Pagano.^24^ R_0_(t) was calculated using previously described methods ^25,26^ assuming a shifted gamma distributed serial interval with a mean of 3.9 days, a standard deviation of 5.8 days based on the estimates of the 2015 wave, which was largely confined to Juba County. We assumed that infection leads to complete immunity over the study period and that 10% of infections were reported as medically attended cholera cases^27^. We fit a quasipoisson model with penalized splines transforming AR7D and lags up to 10 days. The results from this model provide an estimate of the relative R_0_(t) over 10 days following a day with a given value of AR7D compared to an AR7D of 0mm.

We aggregated population movement data to the county level. The origin of returnees immigrating to South Sudan is presented at the country level. Data were most often available at the annual scale (e.g. population movement for 2015/2016 was evenly split between the two years). We calculated the expected number of cholera cases exported by one county to another by multiplying the attack rate for each county by the number of people leaving that area during the same year.

## Results

### First Wave – 2014

After three years without cases, the MoH confirmed a cholera case in Juba County (Central Equatoria State) on April 28, 2014 soon after the start of the rainy season. Case reports increased in Juba County and in the neighboring Eastern Equatoria State (Figures 1 and 2), where CFRs reached as high as 5% (Table 1). Cholera was then reported in the northeastern state of Upper Nile (1,044 cases) where most cases came from an IDP camp outside of Malakal (South Sudan’s second largest city). In total, over the course of 29 weeks, 13 counties reported 6,389 suspected cholera cases (AR=0.05%) and 139 cholera-related deaths (CFR=2.3%, Table 1). Among suspected cases, 25% were children <5 years old and 52% were male. Overall, 430 suspected cases from five different states underwent laboratory confirmation; 44% were confirmed positive for *V. cholerae O1* by culture (Table S1). The last confirmed and suspected case reported in 2014 was on October 27 and on November 8 in Eastern Equatoria State.

**Table 1.**
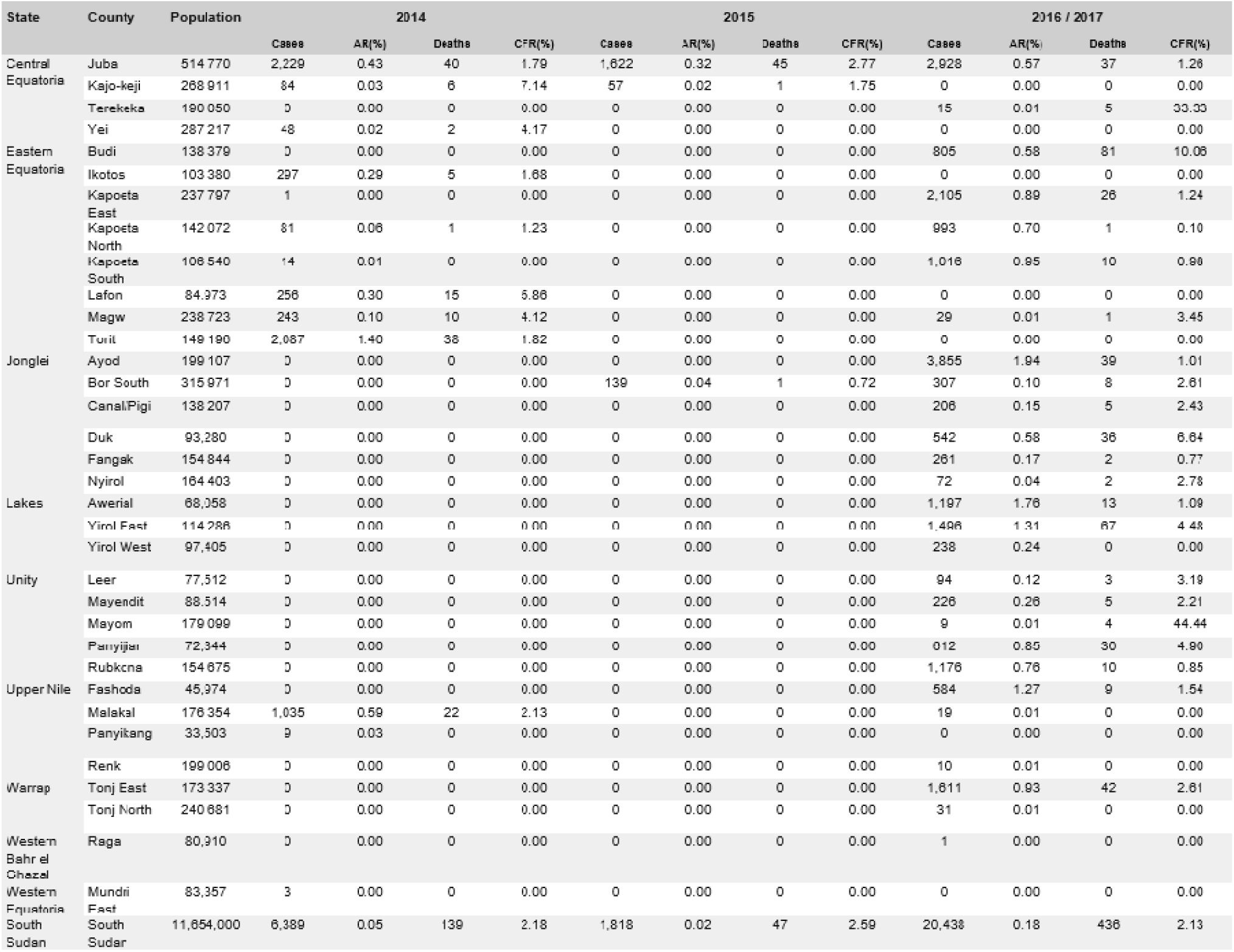
Cholera indicators by county and nationally in South Sudan for each of the three national epidemic waves. AR = Attack Rate. CFR = Case-fatality risk. A nationwide attack rate was calculated for the entire population of South Sudan.

**Figure 1.**
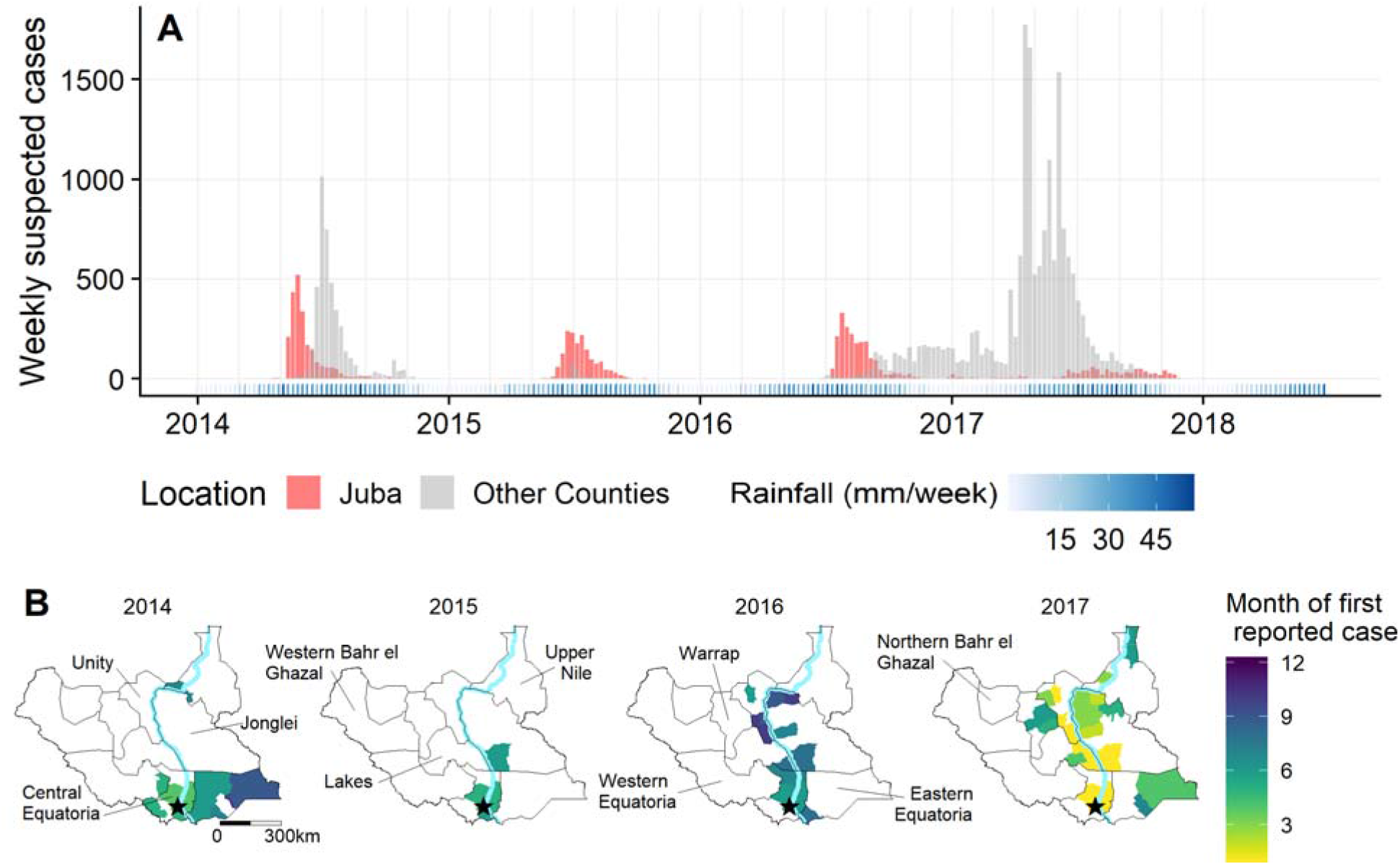
Successive epidemic curves (A) and geographic spread of suspected cholera cases (B) in South Sudan between Jan 1, 2014, and July 1, 2018. Bar heights represent the weekly number suspected cases in Juba County (red) or in other counties (grey). For each year with cholera cases (2014, 2015, 2016 & 2017), county-level maps indicate the month of the first case was reported. States are labeled and borders are in black. Stars represent the capital city of Juba. Light blue line represents the Nile river.

**Figure 2.**
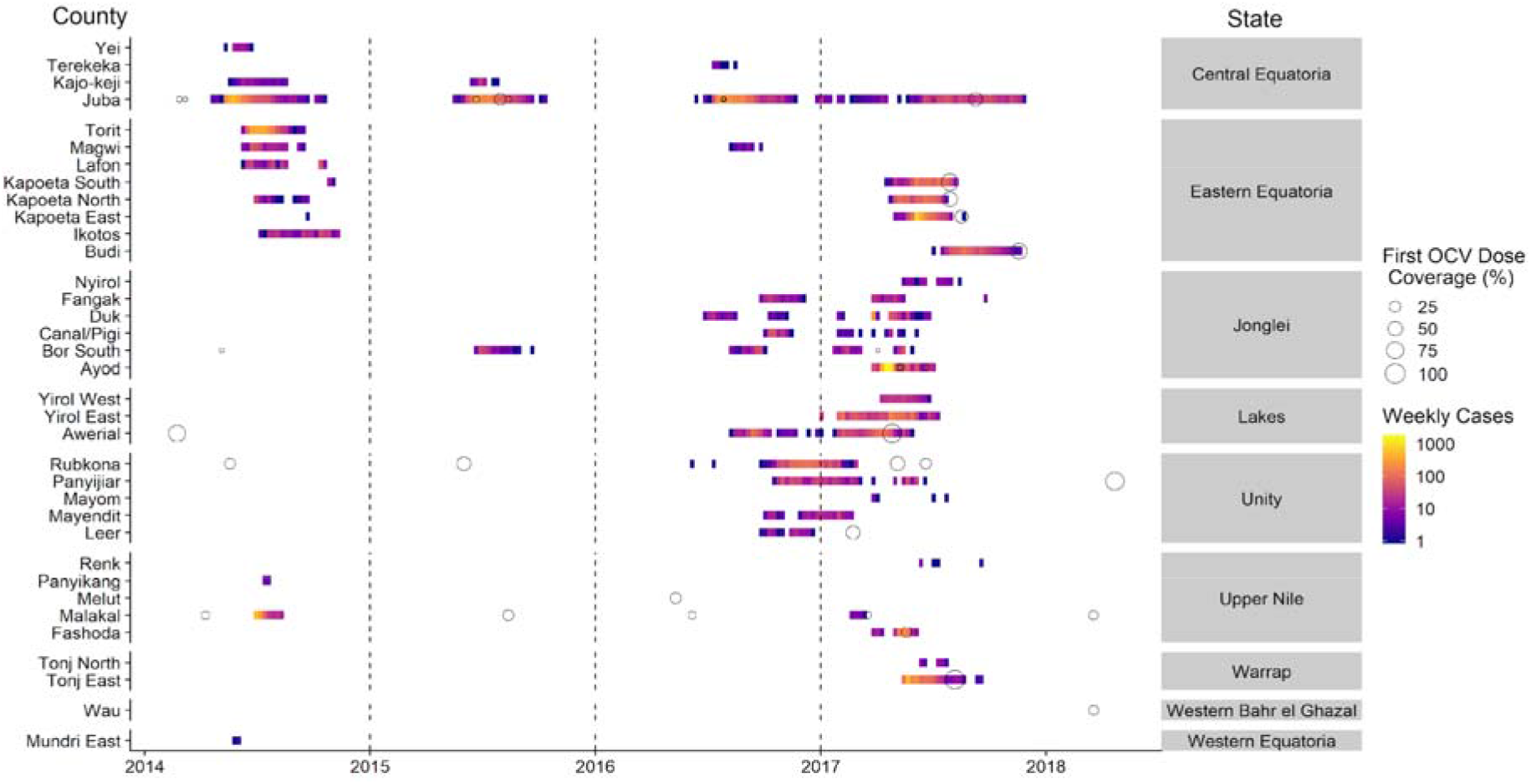
Relative timing of oral cholera vaccine campaigns and county-level cholera outbreaks in South Sudan. Each circle represents a distinct OCV campaign with its size representing the proportion of the county-level population estimated to be vaccinated during the first round (number of doses delivered / estimated population size). Campaigns are marked on the week of the first round, and cases are marked at the week of reported symptom onset for each county.

### Second Wave – 2015

Following a period of 27 weeks with no suspected cases, the first confirmed case of the second wave was reported in Juba County on May 20, 2015. In 2015, over the course of 21 weeks, 1,816 suspected cases (AR=0.02%) and 47 deaths (CFR=2.6%) were reported. Among suspected cases, 18% were <5 years old and 55% were male. Nearly all suspected cases (89%) came from Juba County between May 20 and Oct 15; the remaining 196 cases were reported in two nearby counties (Figure 1B). Out of the 132 samples from suspected cases tested in 2015, 33% were confirmed positive by culture. The last confirmed and suspected cases from the 2015 wave occurred in Juba County on September 11 and October 15, respectively.

### Third Wave – 2016 & 2017

Following a period of 34 weeks with no suspected cases, South Sudan began to experience its largest reported cholera outbreak in recent history, with 20,438 suspected cases (AR=0.18%) and 428 deaths (CFR=2.13%). Among suspected cases, 22% were under five and 48% were male. On June 18, 2016, Juba County reported the first confirmed cholera case of the year within the country, later than both of the previous years. In 2016, 139 of 402 (34.6%) suspected cases were confirmed. Confirmed cases occurred in Central Equatoria State, Eastern Equatoria State, Jonglei State, Lakes State, and Unity State (Table S1). As rainfall decreased from October to December 2016, no additional counties reported cholera.

Through the dry season, counties along the Nile River continued to report suspected cases (Figure 2). Beginning in January, outbreaks were observed in new areas of Lakes State, Unity State, and Jonglei State. Ayod County (Jonglei State) experienced a particularly explosive outbreak with 3,855 suspected cases and 39 deaths occurring between March 27 and July 9, 2017 (>250 cases per week) and 3,276 cases over a 3-week period; many of these cases occurred in cattle camps, where nomadic pastoralists have little to no access to safe water and sanitation ^28^. As the rainy season began in 2017, suspected cases were detected in northeastern (Upper Nile State), southeastern (Eastern Equatoria State), and central (Warrap State) South Sudan. In 2017, 231 of 453 (51.0%) suspected cases across seven states were confirmed by culture. The last confirmed and suspected cases were reported on August 30 and November 29 respectively, with no additional cases reported through the time of revising this manuscript (18-Sept-2020).

### Vaccination Campaigns

In addition to case management, surveillance, and WASH interventions, over 2 million doses of OCV were administered in South Sudan between 2014 and 2018. There were 36 vaccine campaigns (2014: 6, 2015: 6, 2016: 4, 2017: 17, 2018: 3) across 15 of the 34 cholera-affected counties (10 campaigns occurred in Juba County). The population reached during each campaign’s first round (typically out of two) ranged between 1,926 and 206,521. While the administrative coverage (number of doses delivered / estimated population size) of most campaigns in the targeted population was moderate to high (range: 52-119%), the coverage at the county-level was much lower. The median first-dose administrative coverage at the county-level was 18% (IQR: 4-41%) (Figure 2).

Among 25 campaigns conducted reactively (i.e., in response to cases), only 6 occurred before the county-specific peak week of cases. In 2014 and 2015, 4/6 campaigns occurred before the peak as compared to 2/19 campaigns in 2016 and 2017 (Figures 2 & S1). Five campaigns took place after the last suspected case in a county epidemic, all in 2017. All five counties with vaccination campaigns in 2014 or 2015 reported cases during the 2016/2017 wave, though it is unclear if these were in the same sub-county populations. Counties with campaigns occurring after the county-level epidemic peak had, on average, 2.4 (95% CI: 1.8-3.2) times higher attack rates compared to those where vaccine campaigns occurred before the peak, even after adjusting for vaccine coverage and epidemic wave.

### Molecular Analyses

Results from phylogenetic analyses (Figure 3, Figure S2), MLVA, and antibiotic resistance testing (Figure S3) all suggest that the bacteria circulating during the study period were nearly identical *V. cholerae* O1 Inaba strains with the *ctxB1* genotype. Our phylogenetic analysis looked more closely at previously-published T10 lineage sequences from several African countries, including the fourteen sequences from South Sudan within this clade.^14,15^ We added seven recently-published T10 sequences from Uganda in the same phylogenetic tree to better understand the relationship between *V. cholerae* in these neighboring countries.^16^ All fourteen whole genome sequences from South Sudan epidemics between 2014 and 2017,^15^ from patients in Central Equatoria State and Eastern Equatoria State, had extremely similar sequences. Across these fourteen sequences, there are 21 polymorphic sites (∼3% of the diversity observed in the T10 lineage), with no more than 10 SNPs between any two sequences (Figure 3). These sequences all belonged to the T10 lineage, which has circulated in the Democratic Republic of Congo for more than twenty years (Figure S1), was first observed in Uganda in 1998, and is distinct from the T13 lineage recently reported in East and Southern Africa and Yemen.^14,29^ Four sequences from Uganda in 2014–2015 cluster closely with all fourteen sequences from South Sudan.

**Figure 3.**
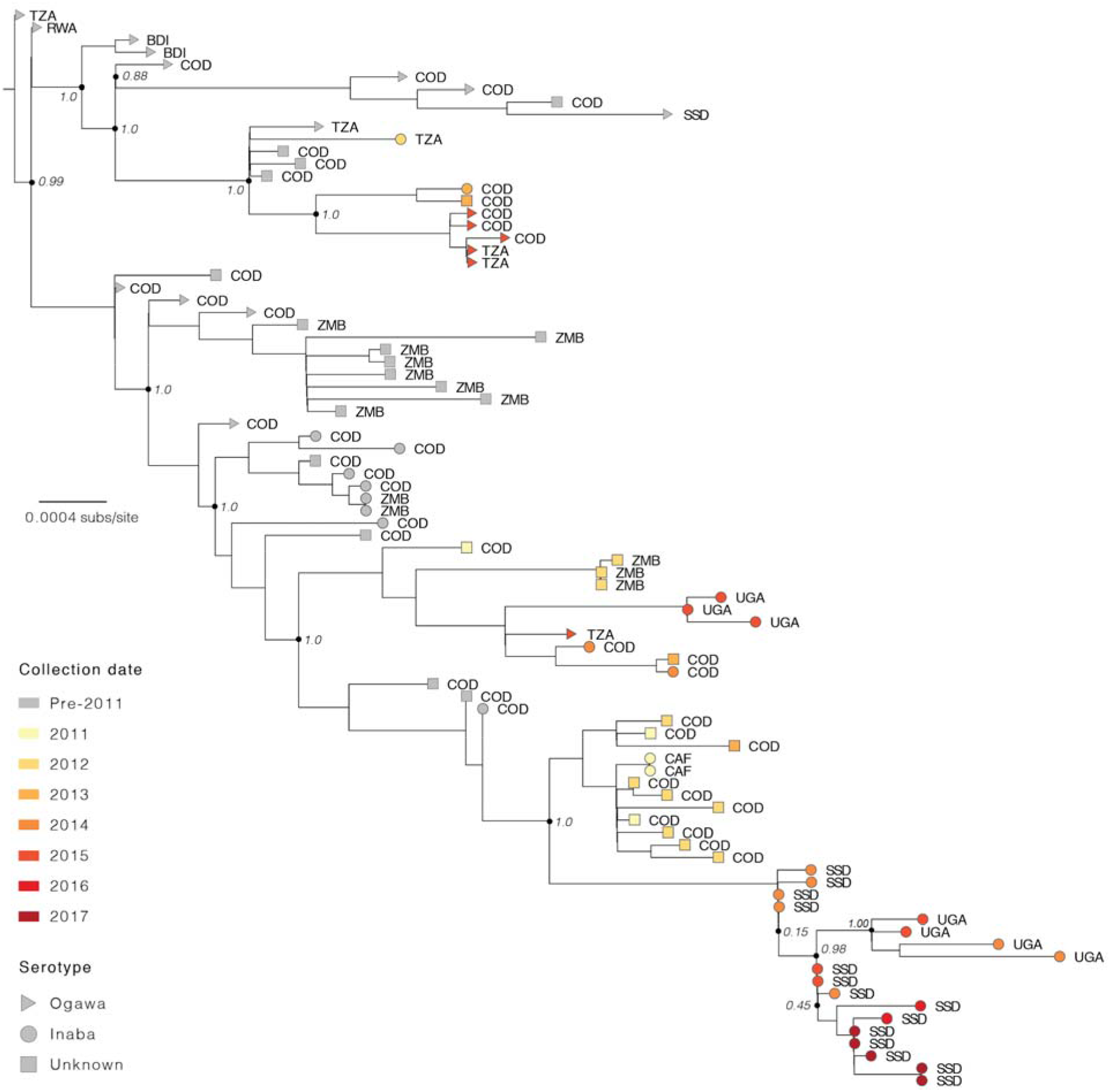
Phylogenetic relatedness of *Vibrio cholerae* O1 El Tor isolates from South Sudan between 2014 and 2017. A subset of T10 lineage isolates published in Weill et al^15,16^, including fourteen isolates from South Sudan, are combined with T10 lineage isolates from Uganda published in Bwire et al.^15,16^Bootstrap values at key nodes are labeled. Country codes: TZA, United Republic of Tanzania; KEN, Kenya; RWA, Rwanda; BDI, Burundi; COD, Democratic Republic of the Congo; ZMB, Zambia; UGA, Uganda; CAF, Central African Republic; SSD, South Sudan.

MLVA analysis on 150 *V. cholerae* isolates collected from 12 different counties in 7 different states in 2014 (46), 2015 (27), 2016 (42), and 2017 (35) showed that all isolates were from the same clonal complex (Figure S3A). Ten isolates collected in Uganda in 2014 also belonged to the same clonal complex. Nearly all isolates had identical antibiotic resistance profiles, including resistance to colistin, nitrofurantoin, polymyxin B, streptomycin, sulfonamides, trimethoprim-sulfamethoxazole (Figure S3B).

### The Role of Precipitation

Over 90% of average annual precipitation during 2010-2018 occurred between March and November, both nationally and in Juba County (Figure S4A). Rainfall in 2014–2017 differed little from that in 2010–2013, when no cholera was reported within the country, despite a 2014– 2016 El Niño-event.^30^ With the exception of 2017, the rainy season preceded the detection of cases every year, with cases in Juba County typically reported in closest proximity (2-10 weeks) to the start of the heavy rains (Figure S4B). In Juba County, we found no significant association between accumulated rainfall over 7 days and the instantaneous basic reproductive number in the subsequent 10 days (Figure S5, S6). Though we did not extend this analysis to all counties, it is noteworthy how this finding contrasts with those from analyses of cholera in in Yemen using similar methods.^21^

### The Role of Population Movement

Between 2014 and 2017, the IOM estimated that over 760,000 individuals moved between counties in South Sudan or immigrated from a neighboring country (Table S2). In 2017, movement increased nearly two-fold (2014: 164,119, 2015: 154,840, 2016: 166,864, 2017: 274,965). Most movement was between counties (553,689) with substantial international migration from Ethiopia (3,025), Democratic Republic of Congo (4,653), Kenya (16,876), Sudan (87,175), and Uganda (95,370).

The movement of people from counties reporting cholera was generally consistent with the year-to-year geographic extent of each wave (Figure 4, Figure S7). In 2014, movement from Juba County, where the first cases were reported, was primarily eastward with little movement north. That same year, cholera was primarily reported in and around Central Equatoria State and in Eastern Equatoria State. In 2015, there was far less movement from Juba County outwards (66% fewer people leaving Juba County than in 2014), and the epidemic remained geographically limited. In 2016 and 2017, movement up and down the Nile River increased dramatically including an increase in movement from Juba County northwards along the river. During this same period, cholera cases were reported from nearly every county along the Nile. In addition to internal migration, more than 110,000 returnees came from Kenya and Uganda, particularly in 2016/17 when both countries reported cholera (Figure S8).

**Figure 4.**
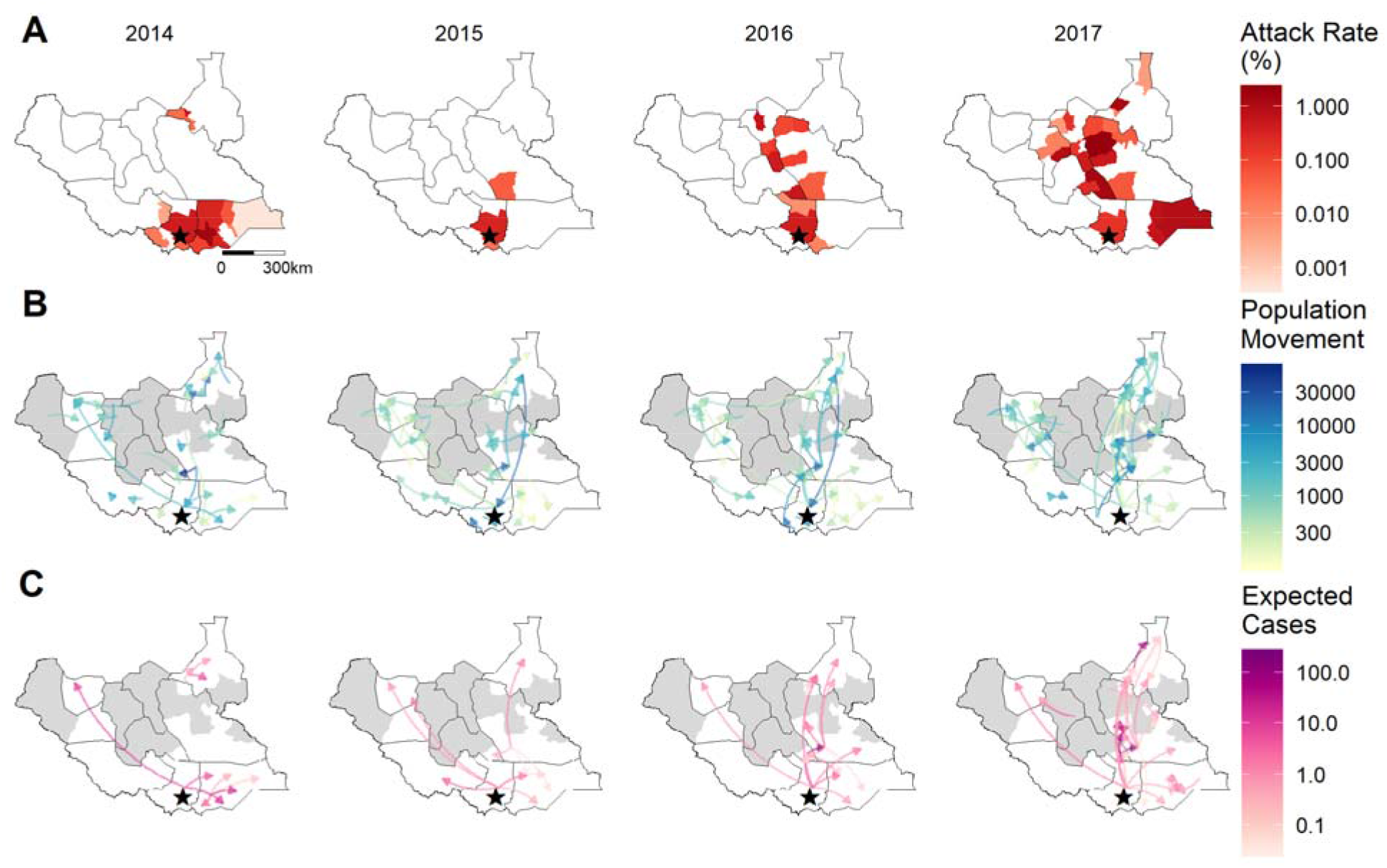
County-level suspected case attack rate and population movement in South Sudan, 2014 - 2017. Red fill (A) indicates the percentage of the population which were suspected cholera cases. Blue arrows (B) represent the number of internally displaced persons and returnees moving between counties. Purple arrows (C) represent the expected number of exported cholera cases estimated by multiplying the attack rate of each county by the number of people leaving that county to another location for the same time period. Higher intensity of population movement and exported cases are indicated by darker colors on the log_10_ scale. Counties in grey were not assessed for arrival of internally displaced persons (IDPs) or returnees. Arrows move to and from county-level centroids.

## Discussion

Between 2014–2017, South Sudan experienced three distinct epidemic waves of cholera, all beginning in Juba County. Genotypic and phenotypic analyses indicate that despite periods of no reported cases in between each wave, all were of the same clonal origin and most closely related to *V. cholerae* O1 circulating in Uganda and Democratic Republic of Congo. Each wave occurred soon after the onset of the rainy season though rainfall was not associated with increased transmission in Juba County. Movements of internally displaced people and returnees were consistent with spatial patterns of cholera, although we could not quantify this association due to the resolution of the data.

Cholera was likely introduced into South Sudan in 2013 or early 2014 through cross-border human population movement.^11^ It remains unclear whether *V. cholerae* O1 remained in South Sudan in between waves through undetected transmission (e.g., insensitive surveillance or asymptomatic carriers), persistence in the environment, or if it was reintroduced. Given the magnitude of population movement between Uganda and South Sudan, reported cases in Uganda during the lull periods in South Sudan, increased surveillance efforts in South Sudan, and the genetic similarity between Ugandan and South Sudanese isolates, repeated cross-border transmission events may be the most plausible explanation. This highlights the need for a regional approach to fighting cholera: efforts to slow an outbreak in one country could be undone by introductions from elsewhere.

For each wave, the first suspected cases in Juba County were detected shortly after the onset of the rainy season. Flooded infrastructure or changes in human behavior around the time of the rainy season likely led to conditions favorable to the propagation of *V. cholerae* O1 in Juba County. Interestingly, increases in rainfall once the rainy season had begun were not associated with increases in transmission in Juba, suggesting rain may have played a role in triggering but not amplifying the outbreaks. In each wave, Juba County was the first county (except narrowly in 2016) to report suspected cases of cholera in the country and likely played an important role in maintaining cholera outbreaks in the country due to the size of the population and the significant population movement in and out of the city (Figure 4, S6).^28^ Juba County had >44% of weeks from 2014-2018 with suspected cholera cases and a mean annual incidence of 2.6 per 1,000, making it a clear ‘burden hotspot,’ as defined by the GTFCC (Figure S9).

Both increased human migration and transmission occurring during the dry season might partially explain why the 2016-2017 outbreak spread more broadly across the country. In July 2016, intense fighting began in Juba County, causing mass population displacement towards swampy areas along the Nile river and to Uganda. At the same time, the number of cases in Juba County was skyrocketing and cases began to be reported from a number of counties adjacent to the river.^31^ Cholera persisted during the dry season only in counties along the river (Figure S10). Though the reason for continued transmission remains unclear, most of these communities had not reported cholera cases in recent years and presumably had low population-level immunity. The large geographic extent at the end of the dry season may have then created conditions where cholera transmission could be easily amplified as heavy rains arrived. Similar dynamics were noted in Yemen where cholera expanded geographically during the dry season with cases exploding when rainfall increased.^21^

The massive use of cholera vaccine (both reactively and proactively) in South Sudan highlights some of the challenges in implementing campaigns, especially in humanitarian settings. Cholera control efforts during these outbreaks was continually hampered by conflicts and limited access to areas with ongoing transmission. Of the 2,068,622 doses used reactively, only 6.4% were part of campaigns initiated before the peak of the epidemic in the target county. Due to the global shortage of vaccine supply, campaigns were often limited in size and rarely covered the full population; only those deemed at the highest risk. Given the low coverage at the county-level (though relatively high coverage in the target population), the spatial scale of the most reliable epidemiologic data collected, and the relatively low testing rate among suspected cases, the impact of vaccination on reducing incidence is unclear. Our simple analyses exploring the relative attack rates in places that had OCV campaigns before and after the peak provide hints of impact on the population-level but efforts to collect more spatially resolved incidence data combined with systematic lab testing of suspected cholera cases will allow for more precise quantification of vaccine impact.

This study comes with a number of limitations. We relied on reports of suspected cases from reporting health structures across the country. However, due to the instability in the region, some health centers may not have reported cases, and some people may not have sought care for even severe disease. Isolates that underwent whole genome sequencing came exclusively from the southern portion of the country and were not geographically representative. However, MLVA analyses and the antibiotic sensitivity testing suggest similarity of strains across the country. While interventions such as chlorination of public water sources, hygiene promotion, and enforcement of sanitation standards in public areas did occur, we were unable to identify datasets to quantify the role of these services. Lastly, the population displacement data were at an annual temporal scale and did not cover the entire country, thus limiting our ability to have precise quantitative estimates of the contribution of movement as shown with cell phone data or theoretical models in other settings. ^32,33^. Despite these limitations, trends of mass migration and spread of cholera were largely consistent.

Cholera has not been reported in South Sudan from November 2017 through the time of last editing this manuscript (19-September-2020), which may be explained by a combination of natural and vaccine-derived immunity, reduced displacement, and perhaps improved sanitary conditions. Estimated water and sanitation indicators remain poor in South Sudan with only 36.7% having access to improved water and 9.9% having access to improved sanitation in 2017.^2^ South Sudan will likely continue to remain at risk of introductions of cholera from other countries in the region followed by large-scale spread. Broad use of oral cholera vaccine combined with local WASH interventions may provide a stopgap over the coming years to block transmission, while much needed investments in water and sanitation are made. However, in South Sudan, peace is likely the first prerequisite to long-lasting cholera prevention.

Through better understanding of how cholera spreads and the complex roles of human movement, climate, and cholera-specific interventions, we can potentially plan for quicker and appropriately targeted cholera control measures in future cholera epidemics in South Sudan and beyond. Such advances in humanitarian emergencies may be critical to achieving progress towards the 2030 cholera goals of reducing cholera as a public health threat.

## Data Availability

The Ministry of Health Republic of South Sudan does not allow the individual record for this dataset to be shared. When possible, we will make available aggregated datasets through a GitHub Repository

## Acknowledgments

We thank Andrea C. Caflisch for his input in interpreting movement data from the IOM. We also thank Daryl Domman for assistance in analyzing sequencing data by sharing alignments and details of masking and alignment methods.

