## Supplementary Appendix for "Successive epidemic waves of cholera in South Sudan, 2014 - 2017"

### Table S1. Laboratory culture confirmation of Vibrio cholerae O1.

Number of samples testing positive for cholera culture from the National Public Health Laboratory in Juba. Three samples in 2017 tested had an unknown geographical state; all were negative.

| State | 2014 | 2015 | 2016 | 2017 |
| --- | --- | --- | --- | --- |
| Central Equatoria | 133/312 (43%) | 39/104 (38%) | 71/200 (36%) | 133/238 (56%) |
| Eastern Equatoria | 39/67 (58%) | 0/6 (0%) | 7/9 (78%) | 14/23 (61%) |
| Jonglei | 1/3 (33%) | 4/17 (24%) | 20/59 (34%) | 20/40 (50%) |
| Lakes | - | 0/1 (0%) | 12/46 (26%) | 33/57 (58%) |
| Unity | - | 0/3 (0%) | 29/76 (38%) | 20/60 (33%) |
| Upper Nile | 16/43 (37%) | - | 0/5 (0%) | 7/24 (29%) |
| Warrap | - | 0/1 (0%) | 0/1 (0%) | 4/8 (50%) |
| Western Bahr el Ghazal | - | - | 0/6 (0%) | - |
| Western Equatoria | 1/5 (20%) | - | - | - |
| South Sudan | 190/430 (44%) | 43/132 (33%) | 139/402 (35%) | 231/453 (51%) |

### Figure S1: Timing of the first round of vaccination campaigns.

Each campaign is represented by a dot. The length of the line is the amount of time between either the peak or last case of a county-level outbreak. Ten campaigns occurred in areas without cholera cases. One campaign in Juba 2015 had a missing date and was excluded from this analysis.

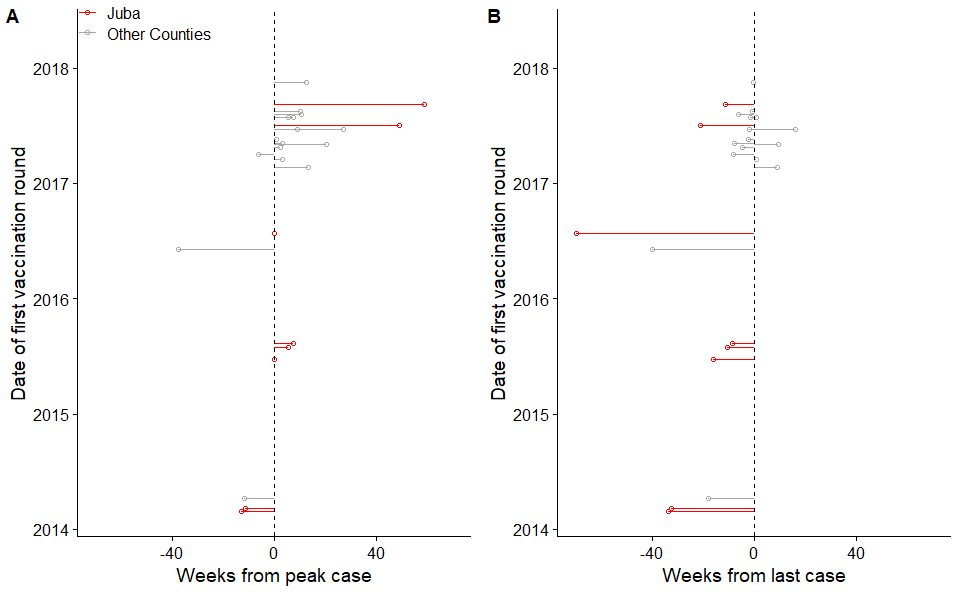

### Figure S2. Maximum likelihood tree of publicly available VC whole core genome sequences.

T10 and T13 lineages, as defined in Weill et al. 2019, are indicated in black; sub-lineage of T10 containing South Sudan isolates (see Fig. 2) indicated in blue; 2014–2017 isolates from South Sudan indicated in orange.

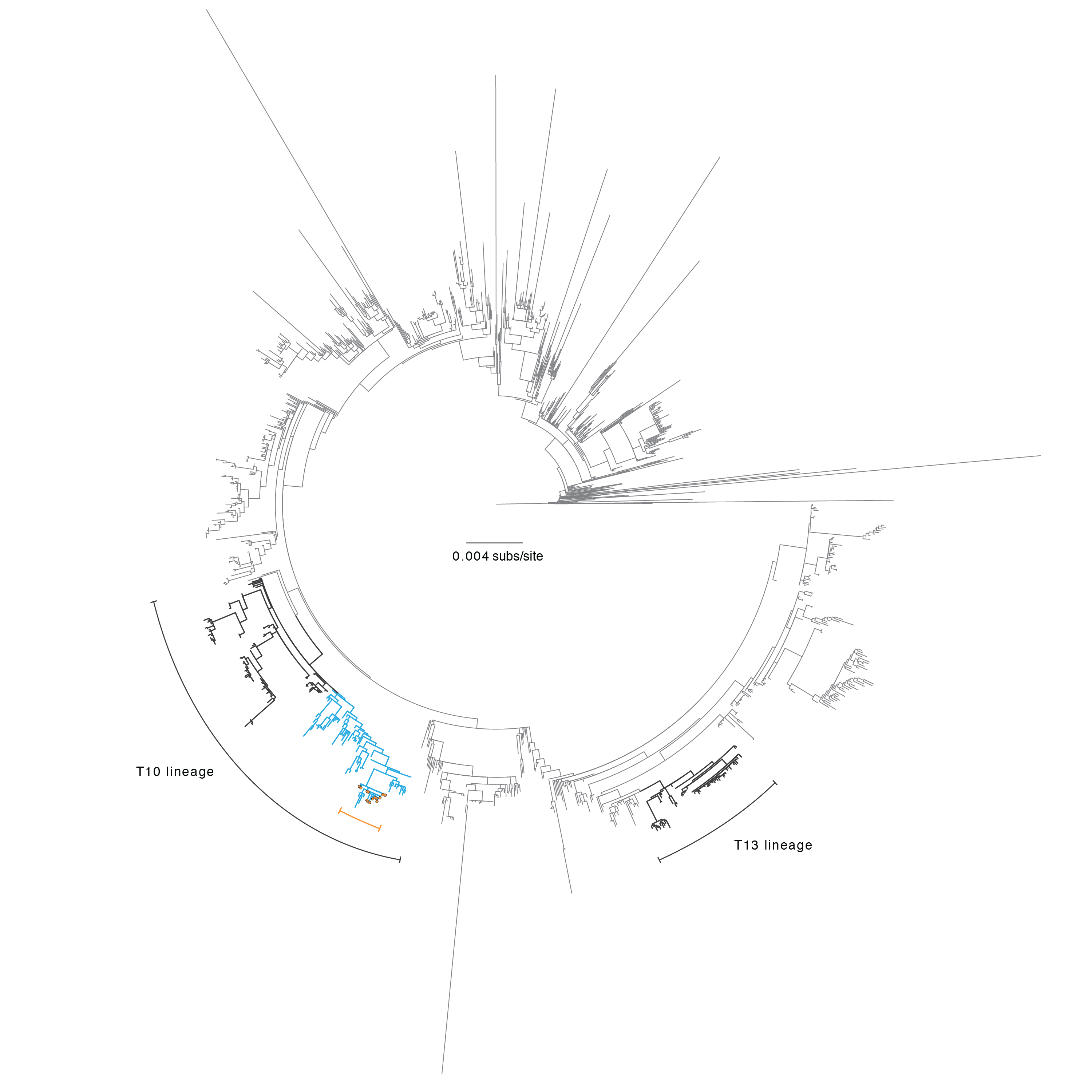

### Figure S3. MLVA and antibiotic sensitivity analyses of V. cholerae isolates from South Sudan

Each row of the heatmaps represents one isolate. The left heatmap shows MLVA data with each column being a single locus. Color represents the number of copies that differ from a reference sequence 10-7-3-7-10-18. The right heatmap shows sensitivity for each sample with each column representing a different antibiotic (sulfonamides, SUL; trimethoprim-sulfamethoxazole, SXT; chloramphenicol, CHL; nitrofurantoin, NIT; erythromycin, ERY; Polymyxin B, PMB; colistin, CST; tetracycline, TET; nalidixic acid, NAL; norflaxacin, NOR; vibriostatic agent, O129; streptomycin, STR). Based on Comité de l’Antibiogramme de la Société Française de Microbiologie 2013 standards for Enterobacteriaceae, nearly all samples were resistant to colistin, nitrofurantoin, polymyxin B, streptomycin, sulfonamides, trimethoprim-sulfamethoxazole and vibriostatic agent O/129 (associated with resistance to trimethoprim) and showed decreased susceptibility to chloramphenicol (Figure S3B); MICs determination with Etest (AB bioMérieux, Solna, Sweden) confirmed resistance of all strains to nalidixic acid as confirmed by detection of one mutation in the gyrA gene (substitution of serine by isoleucine at position 83). Resistance to streptomycin (strAB), sulfonamides (sul2), trimethoprim and O139 vibriostatic agent (dfrA1), and trimethoprim-sulfamethoxazole (sul2 and dfrA1), were reported to be associated with an integrating conjugative element called ICEVchInd5.

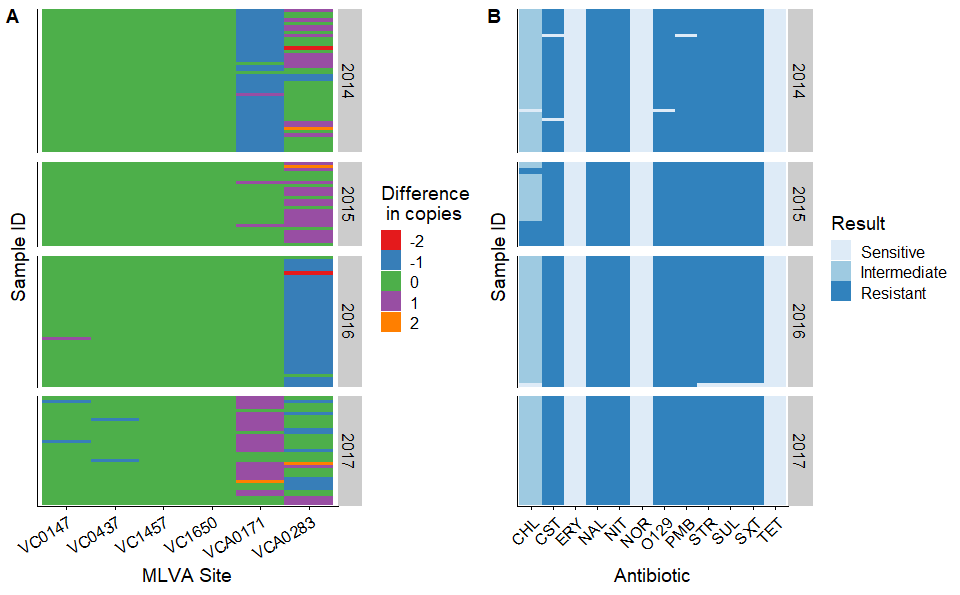

### Figure S4. Temporal association between the start of county-level outbreaks and rainy season, South Sudan, 2014 - 2017

Each point represents a county-level outbreak during a specific year. Color indicates if the outbreak occurred in Juba County (red) or in another county (blue). Solid line represents y=x while the dashed lines are offset by 5 weeks.

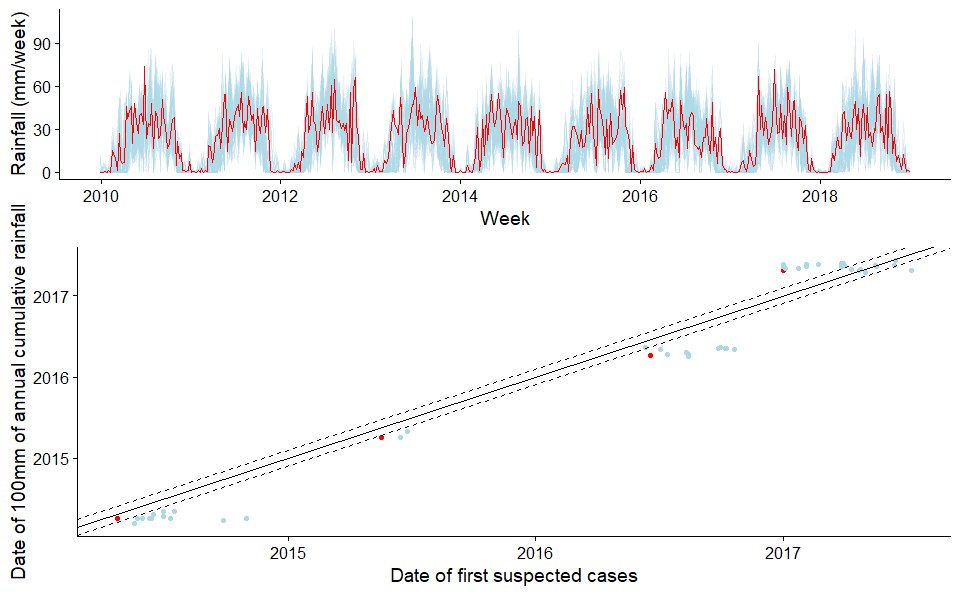

### Equation S1. Distributed lag non-linear model equations

Our approach follows the statistical methods outlined in an analysis of cholera surveillance data from Yemen [1]. We assume that the number of cases reported in Juba, $I(t)$ , on a given day, $t$, follows a Poisson process

$$I(t)\sim Poisson(R(t)\sum_{s=0}^{10} I(t-s)w(s))$$

where $R(t)$ is the instantaneous reproductive number and $w(s)$ is the probability mass function of the serial interval, assuming a shifted (by one day) gamma distributed serial interval with a mean of 3.7 days, a standard deviation of 1.5 days, and a maximum of 10 days [2].

To investigate whether rainfall may have modulated cholera transmission, we accounted for population-level immunity, by estimating the instantaneous basic reproductive number, $R_{0}(t)$. Specifically, we estimated the proportion of the population that is susceptible, $S(t)$, as follows:

$$R(t)=R_{0}(t)\frac{S(t)}{N}$$

$$S(t)=N-\sum_{j=1}^{t-1} I(j)/p$$

where $p$ is the detection probability, assumed to be 10% [3]. Using this set up, we can write out a quasi-poisson regression model equation:

$$log(E[I(t)])=log(R_{0}(t))+log(\sum_{s=1}^{10} I(t-s)w(s))+log(N-\sum_{j=1}^{t-1} I(j)/p)$$

By analyzing the serial interval and susceptibility terms as offset terms, we regressed the instantaneous basic reproductive number on covariates of interest.

$$log(R_{0}(t))=\beta_{0}+\sum_{d=1}^{6} \beta_{dow,d}\delta_{d}(t)+\sum_{k=1}^{3} \beta_{wave,k}\delta_{k}(t)+s(x(t-1),...,x(t-10))$$

$\beta_{0}+\sum_{d=1}^{6} \beta_{dow,d}\delta_{d}(t)$ Specifically, we controlled for the day of the week for potential reporting biases through the week.

$\sum_{k=1}^{3} \beta_{wave,k}\delta_{k}(t)$ We also included a random effect for the wave (Wave 1: 2014, Wave 2: 2015 or Wave 3: 2016/2017) of the epidemic.

Lastly, we included a cross-basis function, $s$, to examine the lagged effects of rainfall using a distributed lag non-linear model (DLNM)

$$s(x(t-1),...,x(t-10))=\sum_{l=1}^{10} f\cdot\omega(x(t-l),l)$$

where $x(t)$ represents the accumulated rainfall over the previous 7 days (AR7D). A maximum lag of 10 days was used, meaning that the AR7D on the previous 10 days was used in the analysis of the instantaneous basic reproductive number on a specific day t. The cross-basis is a sum of bi-dimensional dose-lag-response functions, $f\cdot\omega(x,l)$ each composed of two marginal functions; $f(x)$, the standard rainfall response and $\omega(l)$, the additional lag response function. The marginal functions were both modeled using penalized splines, each with five degrees of freedom. Lastly, we calculated the overall rainfall-response association by summing the risk during the 10-day lag period and centering it to a reference value (AR7D = 0). We fit a generalized additive mixed model (GAMM) using quasi-poisson regression.

### Figure S5. Serial interval and instantaneous basic reproductive number estimates

Results from Bayesian analysis to jointly estimate the serial interval and reproductive are illustrated. Discretized serial intervals used in the analysis are shown and have been normalized to been limited to be over a 10 day period. Estimates of the instantaneous basic reproductive number (estimated using the serial interval from the 2015 cases data) are shown over time.

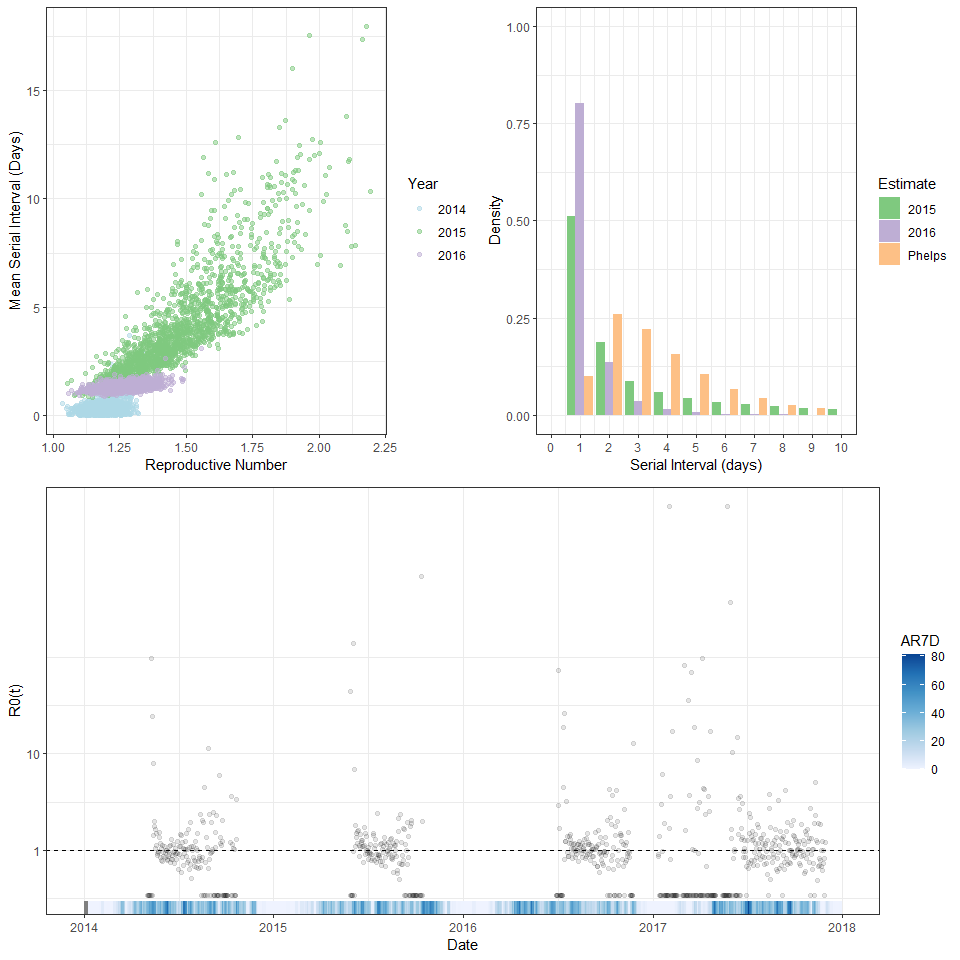

### Figure S6. Analysis of rainfall and cholera transmission using a distributed lag non-linear model

Histogram indicates the number of days with AR7D during which there was cholera was reported. Solid lines are the mean estimate of the relative instantaneous basic reproductive number while shading is the 95% confidence interval.

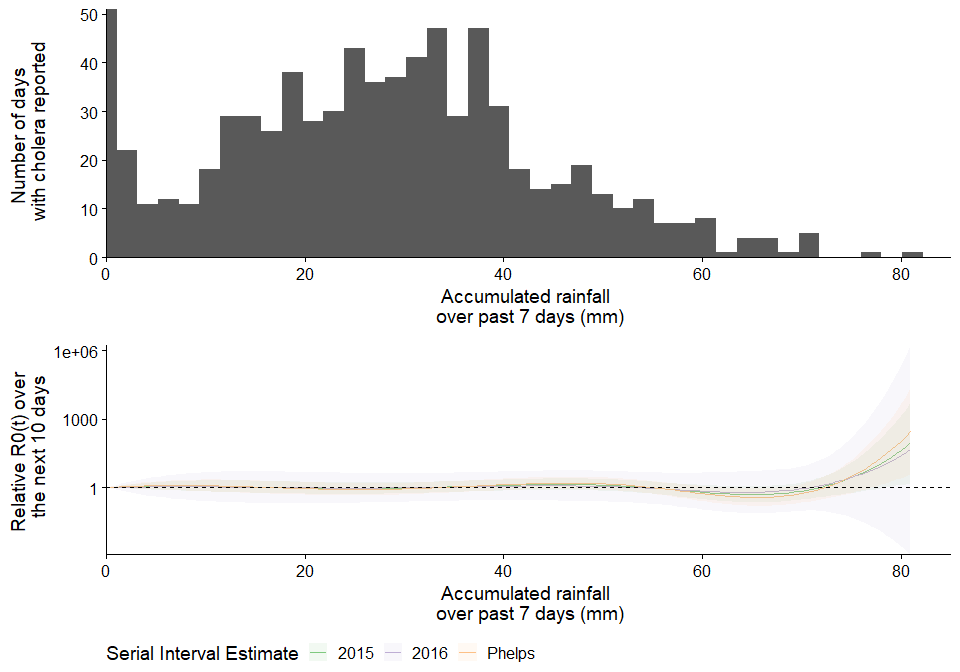

### Table S2. Number of arrivals recorded by the International Office of Migration by country of origin

| Country of Origin | 2014 | 2015 | 2016 | 2017 |
| --- | --- | --- | --- | --- |
| DRC | 0 | 1091 | 658 | 2904 |
| Ethiopia | 0 | 559 | 357 | 2109 |
| Kenya | 0 | 954 | 3291 | 12631 |
| South Sudan | 164119 | 96291 | 121179 | 172100 |
| Sudan | 0 | 27785 | 23779 | 35611 |
| Uganda | 0 | 28160 | 17600 | 49610 |

### Figure S7. Heatmap of domestic population movement between states of South Sudan.

Color is log transformed. If no block exists, no movement was recorded for that particular origin-assessment pair

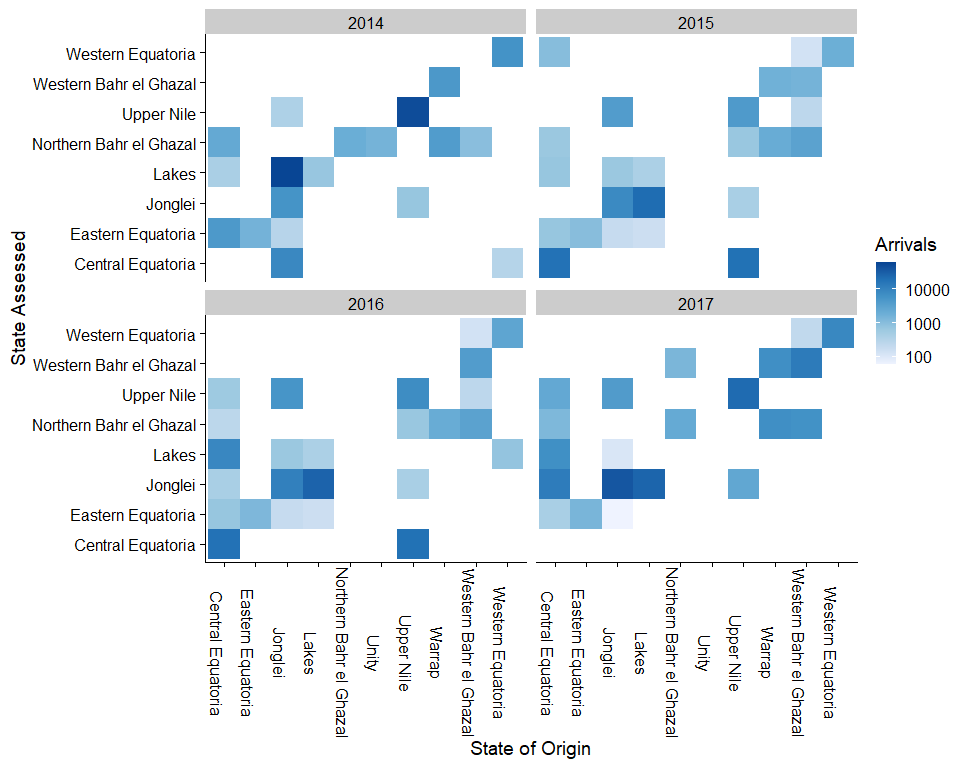

### Figure S8. International cross-border movement of individuals returning to South Sudan (A) and Incidence rates of cholera in neighboring countries (B)

Geographic location is not known below the country level. Incidence rates calculated using WHO reports and population number from (<https://population.un.org/wpp/DataQuery/>). Though Sudan is believed to be endemic for cholera, but has not historically reported it to the WHO. Grey areas were not assessed by the IOM for migration.

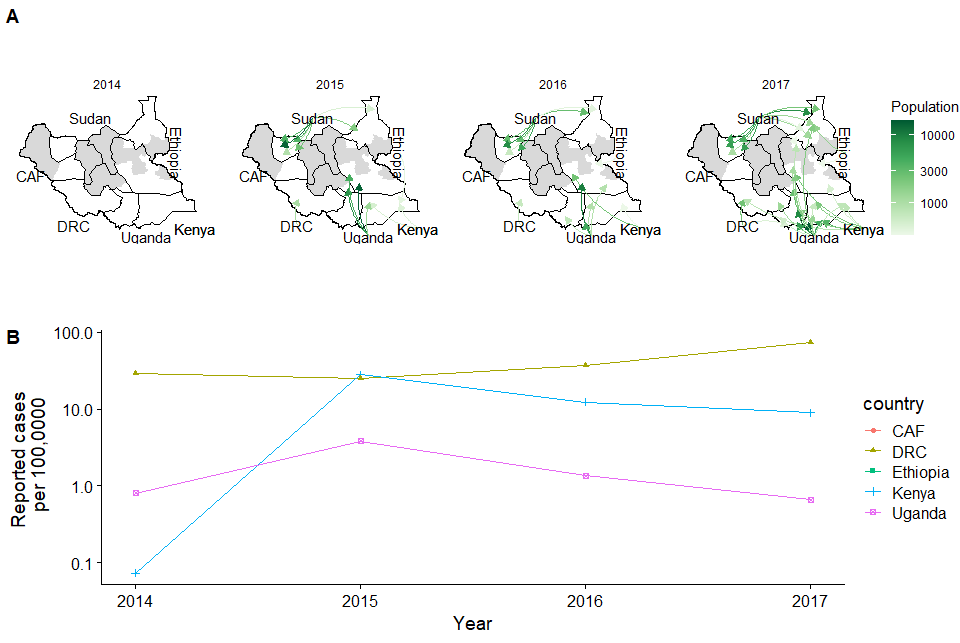

### Figure S9. Analysis of cholera cases in the counties of South Sudan, Jan 2014 - Dec 2018

Incidence and the proportion of weeks with cholera cases are shown for each cholera affected county. Both axes are square-root transformed.

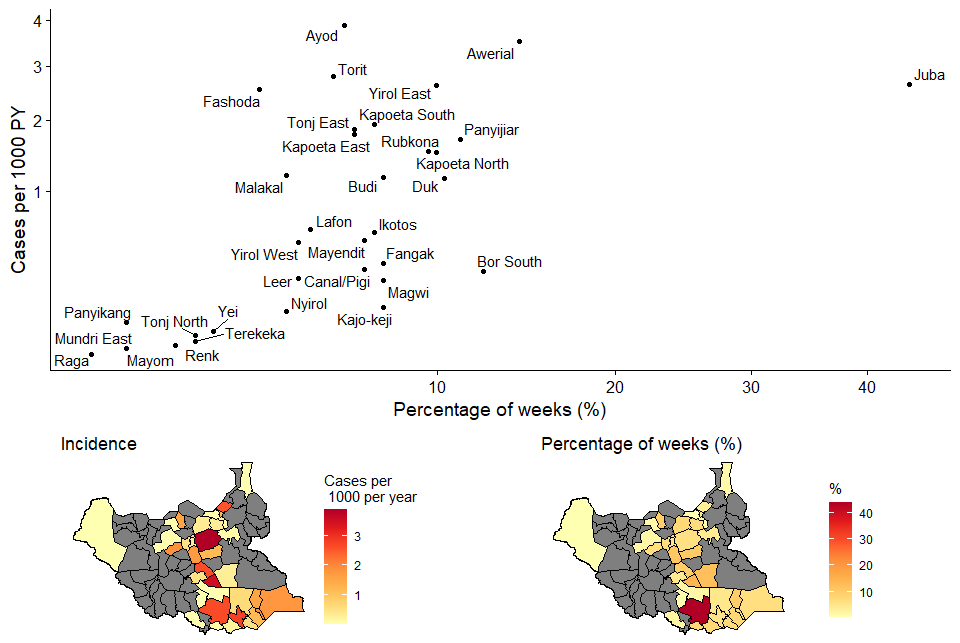

### Figure S10. Monthly geographic distribution of suspected cholera cases in South Sudan, June 2016 - November 2017.

County borders are indicated in black. Light blue line represents the Nile river.

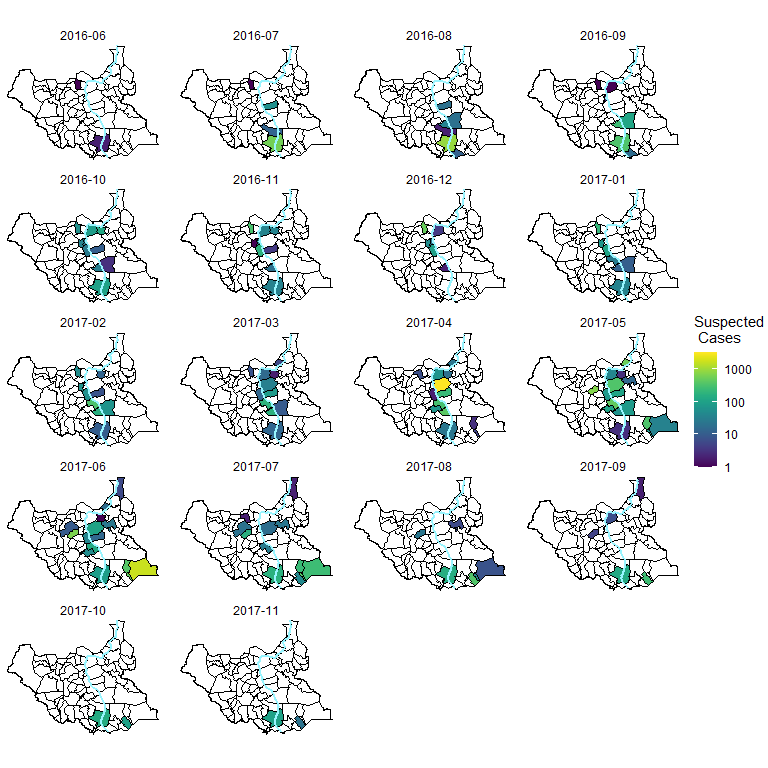
